# Statistical Analysis Plan for the Primary and Selected Secondary Endpoints in the ENHANCED-SPS Study

**DOI:** 10.1101/2023.09.01.23294899

**Authors:** Laura B. Balzer, Jane Kabami, the ENHANCED-SPS Study Team

## Abstract

This document provides the statistical analytic plan (SAP) for the ENHANCED-SPS study, a cluster randomized trial to evaluate the effects of peer-led, multicomponent intervention on viral suppression and other care outcomes among pregnant and breastfeeding women with HIV in South Western Uganda (Clinicaltrials.gov: NCT04122144). The SAP was locked prior to unblinding and effect estimation. This SAP was embargoed until August 31, 2023 when it was submitted to medRxiv.

## 1 Overview

ENHANCED-SPS is a cluster randomized trial designed to evaluate the effectiveness of an enhanced viral load (VL) counseling and standardized peer-mother support (SPS) on viral suppression and other care outcomes among pregnant and breastfeeding women with HIV in South Western Uganda (Clinicaltrials.gov: NCT04122144). The patient-centered intervention was developed using the PRECEDE model of behavioral change and was designed to address the structural barriers and unique needs of the target population. Details of the trial design and procedures can be found in the Study Protocol. This document provides the statistical analysis plan to evaluate the intervention effect on the primary endpoint of viral suppression and selected secondary endpoints, including retention in care, adherence to antiretroviral therapy (ART), and disclosure of HIV status. Analysis plans for qualitative outcomes and other endpoints are available elsewhere. A history of changes to this analysis plan is given in the Appendix.

### 1.1 Study design

ENHANCED-SPS is an Implementation Science study, designed to rigorously evaluate the effects of a multi-component intervention, including viral load counseling and peer support, on care outcomes among pregnant and breastfeeding women with HIV in South Western Uganda. Since the intervention involves clinic-level changes, 14 government-run HIV clinics were randomized to the intervention or the standard-of-care. Randomization was conducted by an independent statistician using a computer-based random number generator and within strata defined by distance, patient burden (HIV prevalence and the number of new HIV diagnoses), and other socioeconomic characteristics (e.g., main occupation of the catchment area of the study clinics).

To facilitate examination of secular trends, retrospective data were collected from all trial clinics (regardless of randomization arm) in the 18-months leading up to intervention initiation. For simplicity, we refer to the pre-intervention phase as “Phase 1” and the intervention phase as “Phase 2”. This document focuses on analyses of Phase 2 data only; analyses of Phase 1 data will be described elsewhere.

The primary objective of this study is to determine whether the intervention improved viral suppression after 12 months of follow-up. Key secondary endpoints include disclosure of HIV status, ART adherence, and retention in care. We also plan to evaluate predictors of these endpoints and describe changes in ART regimen during follow-up.

### 1.2 Description of population and context

Throughout, the target population is pregnant and postpartum women with HIV and accessing care from public health facilities in South Western Uganda. Women with HIV accessing care at the study clinics were eligible for trial enrollment if they were (1) pregnant, previously diagnosed with HIV, and attending the antenatal clinic (ANC) for the first time; (2) pregnant and newly diagnosed with HIV at their first ANC visit; or (3) breastfeeding and newly diagnosed with HIV at the postnatal visit. All eligible and consenting women were consecutively enrolled until the desired clinic-specific sample size was attained (Appendix A.2).

We will provide a participant flow diagram (i.e., a CONSORT diagram) and describe the study population with summary measures of baseline participant characteristics, including age, marital status, enrollment group (pregnant and previously diagnosed; pregnant and newly diagnosed, or breastfeeding and newly diagnosed), ART regimen, and HIV viral suppression status (HIV RNA< 1000 c/mL). Throughout, “baseline” refers to the time of enrollment into Phase 2, when intervention delivery began. We will provide these summary measures overall and by arm. We may also provide these summary measures by clinic.

## 2 Primary endpoint: Virologic suppression after 12 months

For each participant, the primary endpoint is HIV viral suppression after 12 months of follow-up in Phase 2. In the intervention arm, HIV RNA measures (hereafter called “viral loads”) will be obtained through the Ministry of Health (MoH) standard procedures as well as study-specific procedures (GeneXpert point-of-care viral load testing). In the control arm, viral loads will be obtained through MoH standard procedures and accessed via electronic health records (EHR). As a result, the assay used and corresponding lower limit of detection (LLOD) will vary across participants and over time. To be the most inclusive, the primary analysis will define viral suppression as <1000 copies/mL. Pending data availability, we may conduct sensitivity analyses defining viral suppression a <400 copies/mL. Throughout, we will define the “baseline” viral load as the one taken closest to and within 3.5 months of enrollment in Phase 2. Throughout, we will define the “endline” viral load as the one taken closest to and within 3.5 months of 12 months of follow-up in Phase 2.

### 2.1 Missing data

Despite regular viral load monitoring, offered every 6 months or more frequently for pregnant and breastfeeding women, viral load data may be subject to missingness at baseline and at endline. For some women, this missingness may reflect being out of care. For others, this missingness may reflect shortages of viral load assays. Yet, for others, this missingness may reflect gaps in the data system. Given our reliance on routinely collected viral loads and on the EMR system, we anticipate missingness at baseline and endline will be greater in the control arm than the intervention arm.

Our ability to adjust for differential measurement is attenuated by the magnitude of missing outcomes. As a result, we pre-specify the following approach:

1. If 50% or greater of control participants have an endline viral load, we will compare endline viral suppression by arm, as detailed below.
2. If fewer than 50% of control participants have an endline viral load, we will compare the change in viral suppression from baseline to endline in the intervention arm only, as detailed below.

The approach taken for viral suppression, the primary endpoint, will determine the approach taken for the secondary endpoints, specified below.

### 2.2 By arm comparison

If there are sufficient data to support a by arm comparison of endline viral loads (Section 2.1), we will conduct the following analysis. Using targeted minimum loss-based estimation (TMLE),^1–5^ we will compare the proportion of participants with viral suppression after 12 months of follow-up between arms. TMLE accounts for clustering and allows for flexible adjustment for the ways in which participants with measured outcomes could differ from participants missing outcomes. Our adjustment set will consist of study arm, age, enrollment group, and baseline viral suppression status. To minimize the risk of model misspecification bias, we will use Super Learner, an ensemble machine learning method, to combine predictions from generalized linear models, stepwise regression, and the mean.^6^ In the primary analysis, we will use a *<u>Single-stage TMLE</u>* that pools across clinics to estimate and compare the proportion of participants with viral suppression between arms. In a sensitivity analysis, we will use *<u>Two-Stage TMLE</u>* to first estimate the proportion with viral suppression in each clinic, accounting for missing outcomes, and then compare the estimated clinic-specific proportions by arm. In an additional sensitivity analysis, we will conduct an *<u>unadjusted, complete-case analysis</u>*. Specifically, we will restrict to participants with measured outcomes and compare the proportion with viral suppression between arms. All analyses will exclude participants who are missing both baseline and endline viral loads.

All analyses will also account for dependence of participants within clinics. Specifically, for statistical inference, we will aggregate the influence curve to the clinic level,^4,5^ and use the Student’s t-distribution with 14-2=12 degrees of freedom as finite sample approximation to the standard normal distribution.^7^ We test the *null hypothesis* that the ENHANCED-SPS intervention did *not* improve viral suppression, as compared to the standard-of-care, with a one-sided test at the 5% significance level. We will report point estimates and two-sided 95% confidence intervals for the arm-specific endpoints. The primary analysis will be on the difference scale and for the study population (i.e., the sample average treatment effect [SATE]).^2,8^ We will also examine relative effects in secondary analyses.

### 2.3 Within arm comparison

If there are *<u>not</u>* sufficient data to support a by arm comparison of endline viral loads (Section 2.1), we will conduct the following analysis **within the intervention arm only**. In the primary analysis, we will use a *<u>Single-Stage TMLE</u>* to compare the proportion with viral suppression at endline versus baseline, accounting for clustering and for the ways in which participants with measured viral loads could differ from participants with missing viral loads. As described in Section 2.2, we will use Super Learner for flexible estimation; here, our adjustment set will include age and enrollment group. In a sensitivity analysis, we will use *<u>Two-Stage TMLE</u>* to first estimate the proportion with viral suppression at baseline and endline in each clinic (accounting for missing viral loads) and then compare the estimated clinic-specific proportions over time. In an additional sensitivity analysis, we will conduct an *<u>unadjusted, complete-case analysis</u>*. As before, all analyses will exclude participants who are missing both baseline and endline viral loads. We will use the same approach for statistical inference and test the *null hypothesis* that the ENHANCED-SPS intervention did *not* improve viral suppression over time with a one-sided test at the 5% significance level.

### 2.4 Subgroups

To understand effect heterogeneity, we will repeat the above analyses within the following pre-specified baseline subgroups: age group (15-24 years, 25-34 years, >35 years), enrollment group, and marital status. If conducting a by arm analysis, we will also include baseline viral suppression status as a subgroup. Additionally, given the rollout of dolutegravir (DTG) during the study, we will repeat these analyses stratified by women who did and did not switch to DTG during follow-up. Participants with missing data on the variable determining their subgroup classification will be excluded from that analysis.

### 2.5 Predictors

In addition to subgroup analyses, we will report the number and proportion of participants who were not suppressed at baseline and achieved viral suppression at endline – overall and within subgroups. Likewise, we will report the number and proportion of participants who sustained viral suppression (i.e., had viral suppression at both baseline and endline). Additionally, we may conduct the following analyses to understand predictors of viral non-suppression at endline. Within each arm separately, we will estimate unadjusted associations and adjusted associations of viral non-suppression using as predictors: age, enrollment group, marital status, baseline ART regimen, and baseline HIV viral suppression. All analyses will be implemented with the Single-Stage TMLE and yield associations on the relative scale (statistical analogs of the causal risk ratio).

## 3.0 Secondary Endpoints

Evaluation of the intervention effect on the below secondary endpoints will follow from the primary analysis. As detailed in Section 2.1, if we conduct a by arm comparison of viral suppression at endline, then we will compare the secondary endpoints by arm at endline. Alternatively, if we evaluate the change in viral suppression over time in the intervention arm, then we will compare the secondary endpoints between baseline and endline within the intervention arm only.

Key secondary endpoints include

- Disclosure of HIV status, overall and to a partner or spouse
- Adherence to ART
- In-care indicators: an indicator of having at least one clinical visit within {42,60,90} days of {6, 12} months of follow-up
- Time-in-care: the proportion of the 12 month follow-up where the participant is engaged in clinical care. “Out-of-care” time starts 14 days after a missed visit and ends at re-engagement.
- ART covered time: the proportion of the 12 month follow-up where the participant had a full regimen of ART.

Data for the secondary endpoints will be obtained from routine clinical records and study records. For the disclosure and adherence endpoints, we will evaluate the intervention effect using TMLE to account for clustering and for missing outcomes, as described for the primary endpoint (Section 2). For endpoints related to retention in care (i.e., in-care indicators, time-in-care, and ART covered time), we will conduct an unadjusted, complete-case analysis account for clustering. For all secondary endpoints, we will test the null hypothesis of no improvement due to the ENHANCED-SPS intervention with a one-sided test at the 5% significance level. We will also implement analogous subgroup and predictor analyses.

## Data Availability

Not applicable

## Appendix

### A.1 History of changes

This document provides the pre-specified analyses for the primary endpoint of viral suppression and secondary endpoints of disclosure, adherence, and retention in care. For these endpoints, this document supersedes any prior analytic plans (e.g., in the Study Protocol). This document does not cover additional analyses specified in the Study Protocol for pre-post analyses including Phase 1, vertical transmission, costing, or qualitative assessment of facilitators and barriers. Separate statistical analysis plans will be developed for these endpoints.

### A.2 Power calculations

Sample size and power calculations were based on the standard formulas for cluster randomized trials with proportion endpoints.^7^ We expect these calculations to be conservative, because of the precision gained through our pre-specified choice of a one-sided hypothesis test.

We estimated 14 clinics (7 clinics per arm) would provide 80% power to detect at least a 15% absolute increase in viral suppression at 12 months (the primary outcome) from 70% in the standard-of-care, using a two-sided test and assuming a coefficient of variation of *k=0*.*1* (informed by the START trial) and average of 70 participants/clinic. As shown in the following Figure, these calculations are fairly insensitive to the number of participants and viral suppression in the control arm.

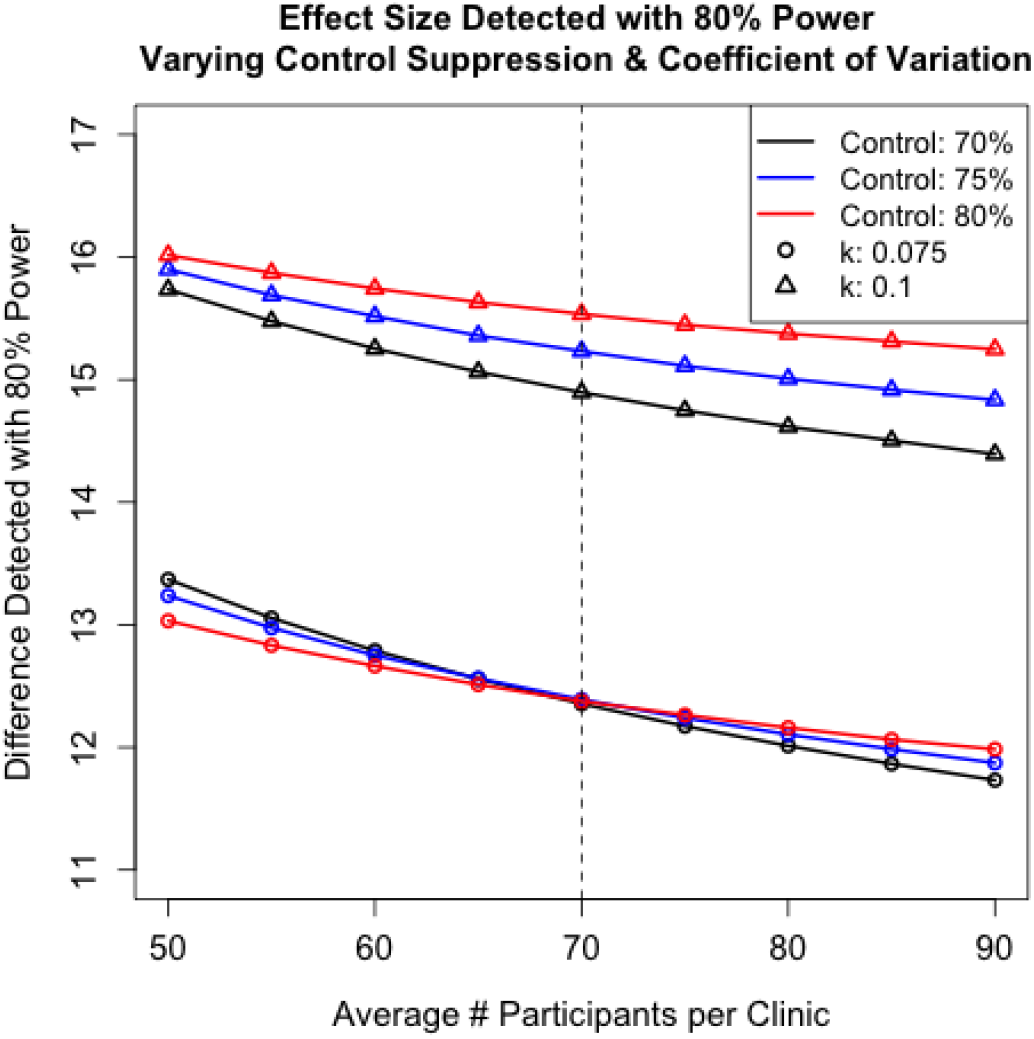

## Notes

### Competing Interest Statement

The authors have declared no competing interest.

### Clinical Trial

NCT04122144

### Funding Statement

Mak-ImS program (FIC NIH Award Number: D43TW010037, 2 D43TW010037)

### Author Declarations

School of Medicine Research and Ethics Committee (SOMREC) of Makerere University (Uganda) and the Uganda National Council for Science and Technology (UNCST) gave ethical approval for this work.

